# mRNA vaccines effectiveness against COVID-19 hospitalizations and deaths in older adults: a cohort study based on data-linkage of national health registries in Portugal

**DOI:** 10.1101/2021.08.27.21262731

**Authors:** Baltazar Nunes, Ana Paula Rodrigues, Irina Kislaya, Camila Cruz, André Peralta-Santos, João Lima, Pedro Pinto Leite, Duarte Sequeira, Carlos Matias Dias, Ausenda Machado

**Affiliations:** Departamento de Epidemiologia, Instituto Nacional de Saúde Doutor Ricardo Jorge, Lisbon, Portugal; Serviços Partilhados do Ministério da Saúde, Lisbon, Portugal; Direção de Serviços de Informação e Análise, Direção-Geral da Saúde, Lisbon, Portugal

## Abstract

**Background:** We used electronic health registries to estimate the mRNA vaccine effectiveness (VE) against COVID-19 hospitalizations and deaths in older adults.

**Methods:** We established a cohort of individuals aged 65 and more years, resident in Portugal mainland through data linkage of eight national health registries. For each outcome, VE was computed as one minus the confounder-adjusted hazard ratio, estimated by time-dependent Cox regression.

**Results:** VE against COVID-19 hospitalization ≥14 days after the second dose was 94% (95%CI 88 to 97) for age-group 65-79 years old (yo) and 82% (95%CI 72 to 89) for ≥80 yo. VE against COVID-19 related deaths ≥ 14 days after second dose was 96% (95%CI 92 to 98) for age-group 65-79 yo and 81% (95%CI 74 to 87), for ≥80 yo individuals. No evidence of VE waning was observed after 98 days of second dose uptake.

**Conclusions:** mRNA vaccine effectiveness was high for the prevention of hospitalizations and deaths in age-group 65-79 yo and ≥80 yo with a complete vaccination scheme, even after 98 days of second dose uptake.

## Introduction

Vaccination is essential to reduce COVID-19 burden and its complications. The first published observational studies showed that the mRNA (Pfizer-BioNTech and Moderna) vaccines were highly effective in preventing SARS-CoV-2 infections and severe outcomes in older adults, even with partial vaccination [1,2]. Vaccine effectiveness (VE) against hospital admissions and COVID-19 related mortality in elderly populations varied between 43-64% for the partial vaccination and 94-97% for the completed vaccination scheme in December 2020-April 2021 [1,3–5].

Recent research showed a decrease in VE against the Delta SARS-CoV-2 variant of concern (VOC) and some evidence of waning immunity for symptomatic infections in the general population [6,7], but sustained VE estimates against hospitalizations [7,8]. However, most studies of VE against severe forms of COVID-19 had a limited follow-up period after complete vaccination [1,4,5], estimated VE of a single dose [2,3] or focused on the general population [7,8] and few considered COVID-19 related mortality outcome and high-risk groups. Understanding VE against outcomes of various severity levels in diverse epidemiological contexts for high-risk groups is important to inform public health measures.

This study aimed to estimate mRNA (Pfizer-BioNTech and Moderna) COVID-19 vaccines effectiveness against hospitalizations and COVID-19 related deaths in Portuguese adults aged 65 years old or more in February-August 2021.

## Methods

### Study settings

We developed a cohort study, based on linkage of electronic health registries. The target population included individuals aged 65 or more years old (yo) resident in private households in Portugal mainland eligible for vaccination.

We excluded from the analysis individuals with improbable ages (more than 110 yo), institutionalized, residents in autonomous regions of Madeira and Azores and with previous SARS-CoV-2 infection. Additionally, to improve completeness and quality of the health records, we included only the subset of National Health Service frequent users[9] for 65-79 yo. For the ≥80 yo cohort, data analysis was restricted to those with at least one vaccine uptake in the last five years for influenza or pneumococcal vaccines.

The study period was defined based on the vaccination campaign roll-out calendar, starting on the 2nd of February 2021 for ≥80 yo cohort and on the 30th of March 2021 for the 65-79 yo cohort, up to the date of the last observed event for each outcome (Table S1).

### Data sources

Eight national electronic health registries were used in this study, including the National Health Service User (NHSU) databaset, the vaccination registry (VACINAS), the National Information System for Epidemiologic Surveillance (BI-SINAVE), the National Death Registry (SICO), the Primary Care Information System (SIM@SNS), the Primary Care Clinical Monitoring System of COVID-19 Patients in Home Isolation (BI-TRACE COVID-19), the National Database of Medicine and Treatment Prescriptions (PEM) and the national database of hospital discharges (BIMIH). All the data from these registries was centralized in one analytic system (CovidAnalytics).

Data extraction and linkage was performed on 13^th^ August 2021 by the Shared Services of the Ministry of Health in accordance with legal and ethical requirements. All data were anonymized prior to statistical analysis by the research team. The study protocol was approved by the Data Protection Officer and the Ethical Committee of the Instituto Nacional de Saúde Doutor Ricardo Jorge (INSA)

### Variables

#### Outcome

A COVID-19 hospitalization was defined as admission for at least 24h with COVID-19 as the primary diagnosis (ICD10 code U07.1) [10], and a previous positive reverse transcription polymerase chain reaction (RT-PCR) test. All-cause death with positive RT-PCR test occurred within previous 30 days were considered COVID-related death [4].

#### Exposure

Exposure to mRNA vaccine was categorized into three levels: unvaccinated (no dose registered), partially vaccinated (14 days after the first dose or less than 14 days after the 2nd dose) and with complete vaccination (14 days after the 2nd dose uptake). To evaluate the hypothesis of vaccine effectiveness waning, time starting at 14 days after the second dose was stratified in 28-day intervals.

#### Confounding

Age groups, sex, municipality level European Deprivation Index (EDI) quintile [11], health region, number of chronic conditions (among anemia, asthma, cancer, cardiac disease, stroke, dementia, diabetes, hypertension, chronic liver disease, neuromuscular disease, renal disease, rheumatologic disease pulmonary disease, obesity, immunodeficiency, tuberculosis), number of laboratory SARS-CoV-2 tests during 2021, previous influenza or pneumococcal vaccine uptake in the last 3 years were considered as potential confounders.

### Statistical methods

We compared individual characteristics at baseline by vaccine exposure status and estimated the COVID-19 hospitalization and death rates per 1000 person-years for unvaccinated exposure and mRNA vaccine exposure period according to the number of doses. Individuals vaccinated with other vaccine brands contributed to unvaccinated exposure prior to vaccination. Given the different observational periods, we estimated VE separately for two age-group cohorts: 65-79 and ≥80 yo. VE was computed as one minus the confounder-adjusted hazard ratio, for each outcome, estimated by time-dependent Cox regression [2] with time dependent vaccine exposure, adjusted for confounding using 7-day periods as strata. Statistical analysis was performed in R Computing Environment, version 4.0.5.

## Results

### Participants

We enrolled 1,409,831 persons aged 65-79 yo and 470,520 aged ≥80 yo in the study (Figure S1, S2). Among 65-79 yo, 641,119 (45.5%) received the Pfizer-BioNTec, 112,132 (8.0%) the Moderna and 531,342 (37.7%) other vaccines, while 125,338 (8.9%) remained unvaccinated. In the ≥80 yo cohort 5.7% were unvaccinated whereas 80.4%, 11.8% and 2.1% received at least one dose of Pfizer-BioNTec, Moderna or other vaccines, respectively. We present the roll-out of vaccine coverage in supplementary files (Figure S3).

The most relevant differences between unvaccinated and vaccinated individuals were observed in their distribution by health region, the number of chronic conditions and previous vaccination for influenza or pneumococcal in the last 3 years (Table 1 & 2). The exposure time to the second dose varied between the two age cohorts, with a median of 78 days (IQR 71 to 94) for the 65-79 yo and 125 days (IQR 112 to 145) for the ≥80 yo cohort (Table S2).

**Table 1:**
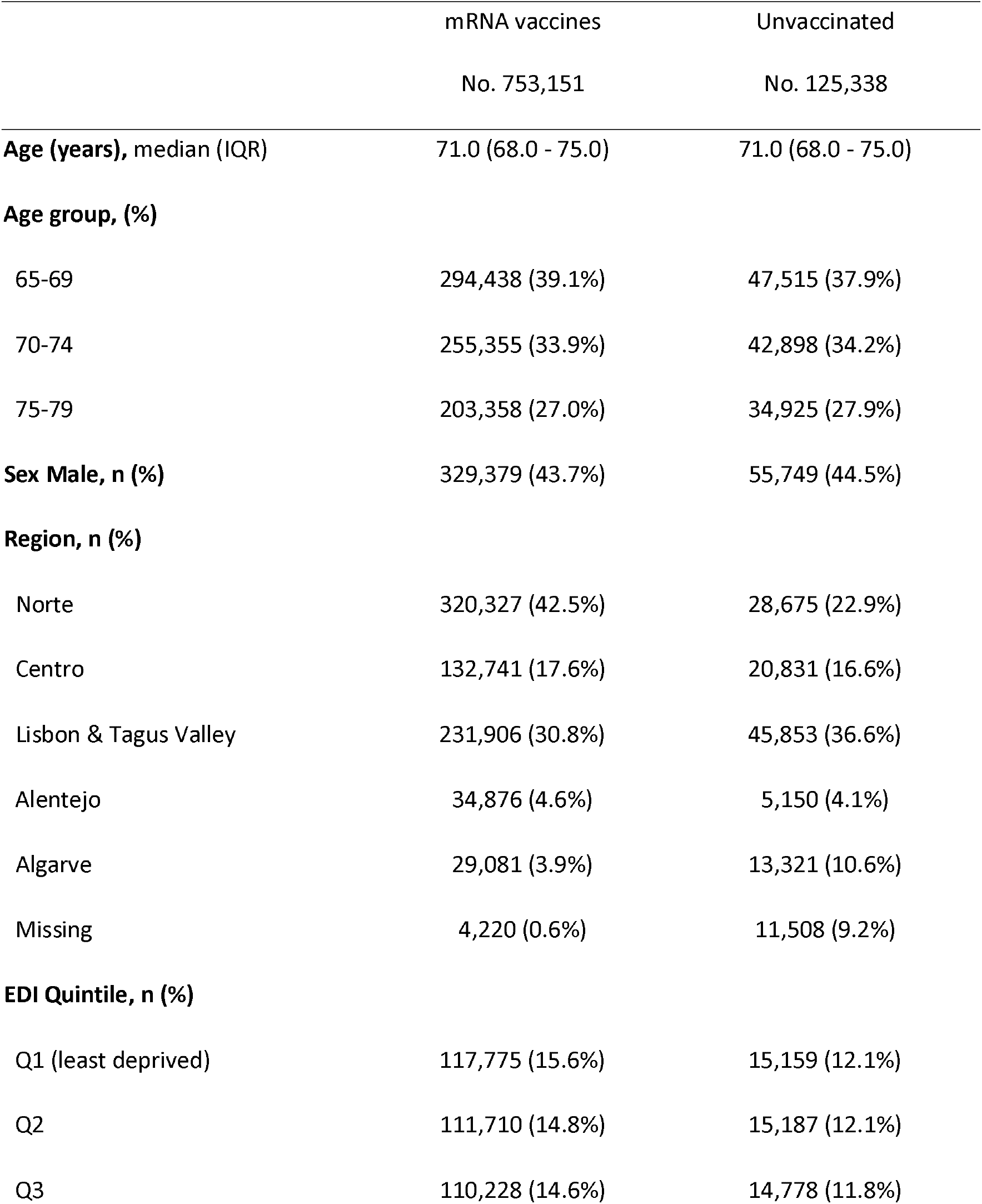
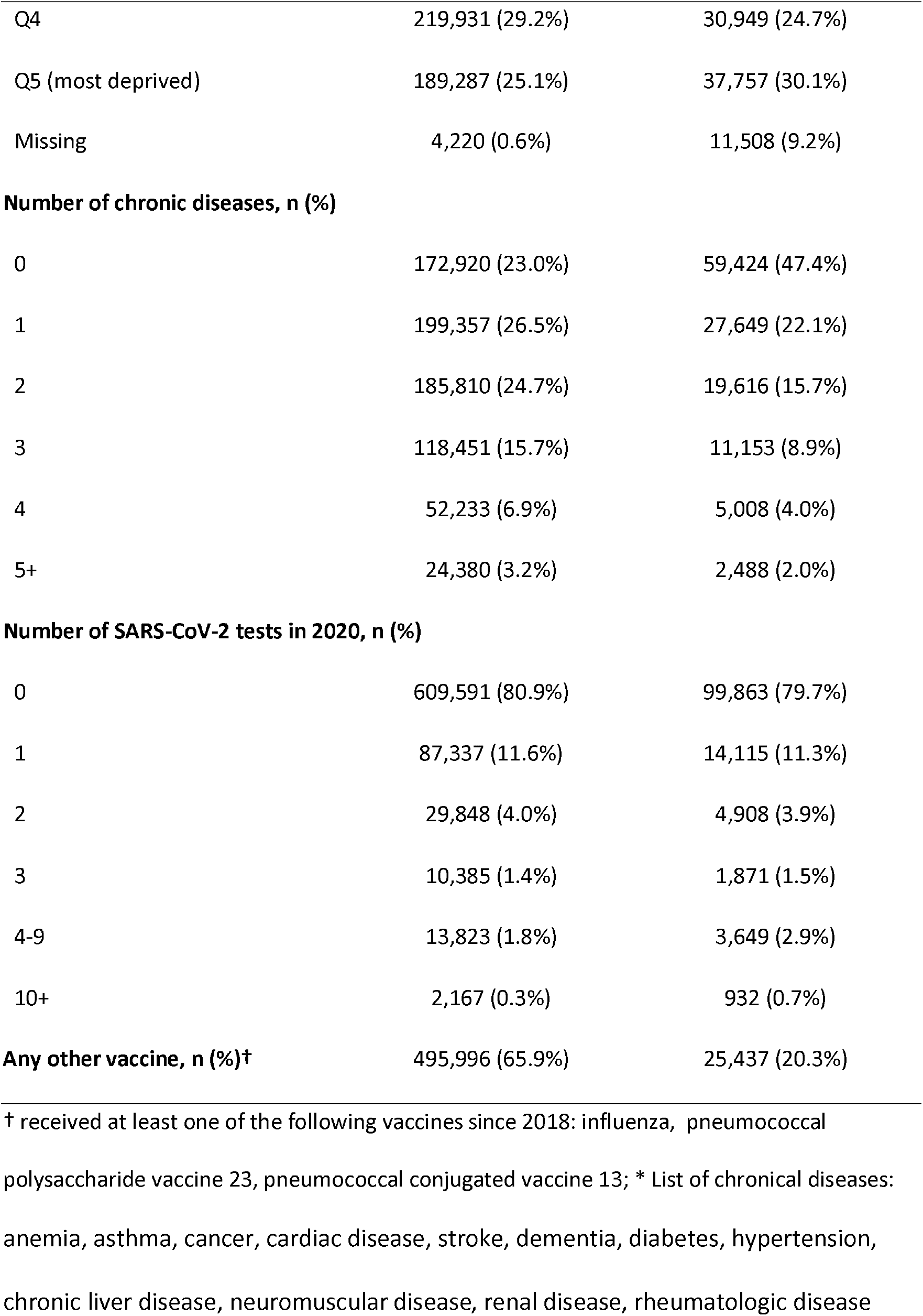
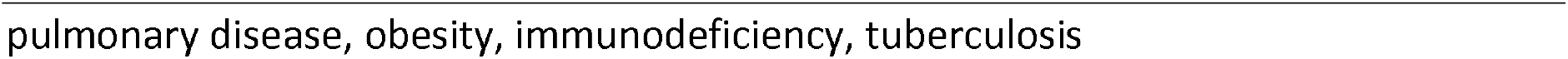
Distribution of 65 to 79 years of age cohort individuals by age group, sex, health region, municipality level European Deprivation index, number of chronic diseases*, number of SARS-CoV-2 tests performed in 2021, influenza or pneumococal vaccine uptake in the last three years.

**Table 2:** Distribution of 80 years and more cohort individuals by age group, sex, health region, municipality level European Deprivation index, number of chronic diseases*, number of SARS-CoV-2 tests performed in 2021, influenza or pneumococal vaccine uptake in the last five years.

|  | mRNA | Unvaccinated |
| --- | --- | --- |
|  | No. 433,878 | No. 26,942 |
| Age (years), median (IQR) | 84.0 (82.0 - 88.0) | 86.0 (83.0 - 90.0) |
| **Age group, (%)** |  |  |
| 80-84 | 222,087 (51.2%) | 10,342 (38.4%) |
| 85-89 | 144,989 (33.4%) | 9,197 (34.1%) |
| 90-94 | 54,046 (12.5%) | 5,301 (19.7%) |
| 95+ | 12,756 (2.9%) | 2,102 (7.8%) |
| **Sex Male, n (%)** | 176,386 (40.7%) | 9,628 (35.7%) |
| **Region, n (%)** |  |  |
| Norte | 159,051 (36.7%) | 8,874 (32.9%) |
| Centro | 91,672 (21.1%) | 5,145 (19.1%) |
| Lisbon & Tagus Valley | 141,890 (32.7%) | 9,284 (34.5%) |
| Alentejo | 24,013 (5.5%) | 1,243 (4.6%) |
| Algarve | 15,778 (3.6%) | 1,594 (5.9%) |
| Missing | 1,474 (0.3%) | 802 (3.0%) |
| **EDI Quintile, n (%)** |  |  |
| Q1 (least deprived) | 75,836 (17.5%) | 4,273 (15.9%) |
| Q2 | 67,922 (15.7%) | 3,759 (14.0%) |
| Q3 | 65,827 (15.2%) | 3,981 (14.8%) |
| Q4 | 120,327 (27.7%) | 7,200 (26.7%) |
|  | No. 433,878 | No. 26,942 |
| Q5 (most deprived) | 102,492 (23.6%) | 6,927 (25.7%) |
| Missing | 1,474 (0.3%) | 802 (3.0%) |
| **Number of chronic diseases, n (%)** |  |  |
| 0 | 45,350 (10.5%) | 9,325 (34.6%) |
| 1 | 84,118 (19.4%) | 4,279 (15.9%) |
| 2 | 112,888 (26.0%) | 4,940 (18.3%) |
| 3 | 96,043 (22.1%) | 4,249 (15.8%) |
| 4 | 56,889 (13.1%) | 2,393 (8.9%) |
| 5+ | 38,590 (8.9%) | 1,756 (6.5%) |
| **Number of SARS-CoV-2 tests in 2021, n (%)** |  |  |
| 0 | 338,916 (78.1%) | 17,503 (65.0%) |
| 1 | 48,115 (11.1%) | 3,665 (13.6%) |
| 2 | 19,427 (4.5%) | 1,976 (7.3%) |
| 3 | 9,373 (2.2%) | 1,135 (4.2%) |
| 4-9 | 16,176 (3.7%) | 2,355 (8.7%) |
| 10+ | 1,871 (0.4%) | 308 (1.1%) |
| **Any other vaccine, n (%)†** | **418,873 (96.5%)** | **22,518 (83.6%)** |

† received at least one of the following vaccines since 2018: influenza, Pneumococcal polysaccharide vaccine 23, pneumococcal conjugated vaccine 13; \* List of chronic diseases: anemia, asthma, cancer, cardiac disease, stroke, dementia, diabetes, hypertension, chronic liver disease, neuromuscular disease, renal disease, rheumatologic disease, pulmonary disease, obesity, immunodeficiency, tuberculosis

During the observation period, a total of 218 COVID-19 hospital admissions and 121 deaths were registered for the 65-79 yo cohort whereas among the ≥80 yo 836 cases were hospitalized with primary COVID-19 diagnosis and 701 died (Figure S4, S5).

### Vaccine effectiveness against hospitalizations with primary diagnosis of COVID-19

For the 65-79 yo, adjusted mRNA VE against COVID-19 hospitalization was 78% (95% CI 61 to 87) for partial vaccination and 94% (95% CI 88 to 97) for complete vaccination scheme (Table3). For ≥80 yo cohort, we observed lower VE estimates for hospitalization, 55% (95% CI 36 to 69) for partial and 82% (95% CI 72 to 89) for complete vaccination, respectively (Table 4).

**Table 3:**
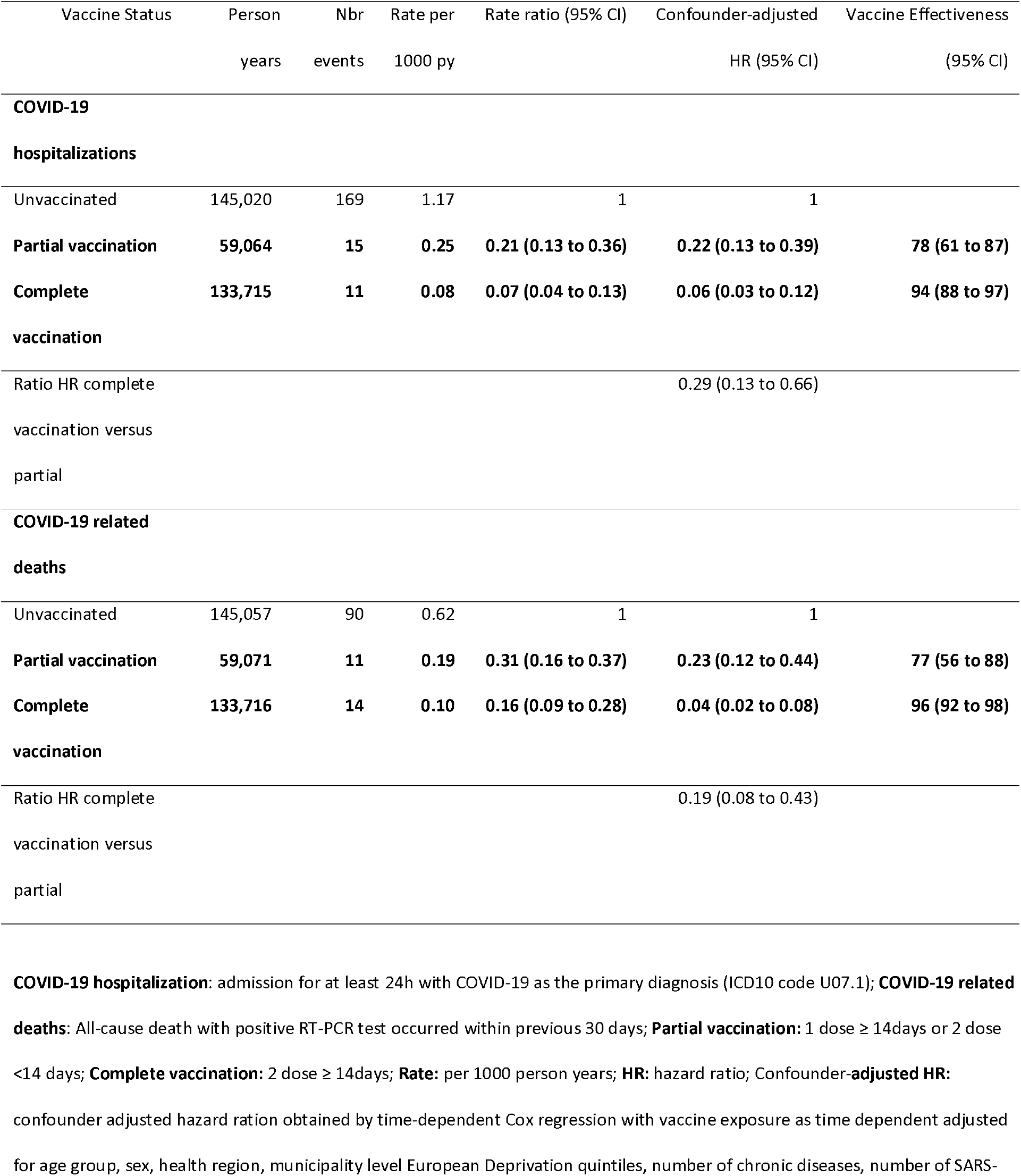
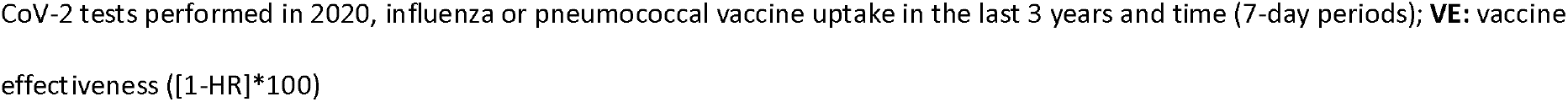
COVID-19 hospitalizations and COVID-19 related deaths, crude incidence rate (1000 person years), confounder-adjusted hazard ratios and vaccine effectiveness (%) by number **of mRNA doses**, for **individuals aged 65-79 years old**.

| Vaccine Status | Person<br>years | Nbr<br>events | Rate per<br>1000 py | Rate ratio (95% CI) | Confounder-adjusted<br>HR (95% CI) | Vaccine Effectiveness<br>(95% CI) |
| --- | --- | --- | --- | --- | --- | --- |
| **COVID-19** |  |  |  |  |  |  |
| **hospitalizations** |  |  |  |  |  |  |
| Unvaccinated | 145,020 | 169 | 1.17 | 1 | 1 |  |
| **Partial vaccination** | **59,064** | **15** | **0.25** | **0.21 (0.13 to 0.36)** | **0.22 (0.13 to 0.39)** | **78 (61 to 87)** |
| **Complete** | **133,715** | **11** | **0.08** | **0.07 (0.04 to 0.13)** | **0.06 (0.03 to 0.12)** | **94 (88 to 97)** |
| **vaccination** |  |  |  |  |  |  |
| Ratio HR complete |  |  |  |  | 0.29 (0.13 to 0.66) |  |
| vaccination versus |  |  |  |  |  |  |
| partial |  |  |  |  |  |  |
| **COVID-19 related** |  |  |  |  |  |  |
| **deaths** |  |  |  |  |  |  |
| Unvaccinated | 145,057 | 90 | 0.62 | 1 | 1 |  |
| **Partial vaccination** | **59,071** | **11** | **0.19** | **0.31 (0.16 to 0.37)** | **0.23 (0.12 to 0.44)** | **77 (56 to 88)** |
| **Complete** | **133,716** | **14** | **0.10** | **0.16 (0.09 to 0.28)** | **0.04 (0.02 to 0.08)** | **96 (92 to 98)** |
| **vaccination** |  |  |  |  |  |  |
| Ratio HR complete |  |  |  |  | 0.19 (0.08 to 0.43) |  |
| vaccination versus |  |  |  |  |  |  |
| partial |  |  |  |  |  |  |

**Table 4.**
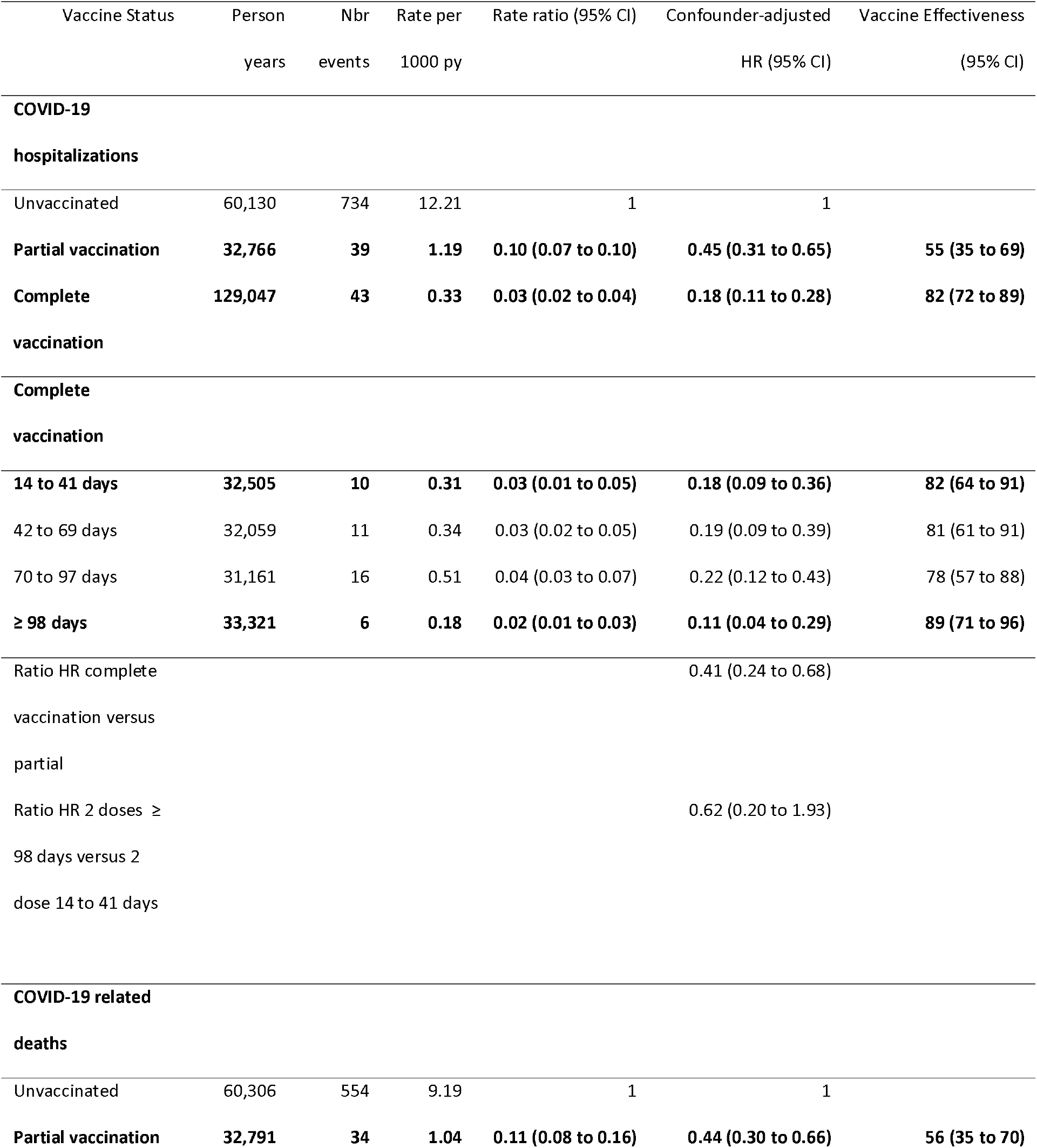
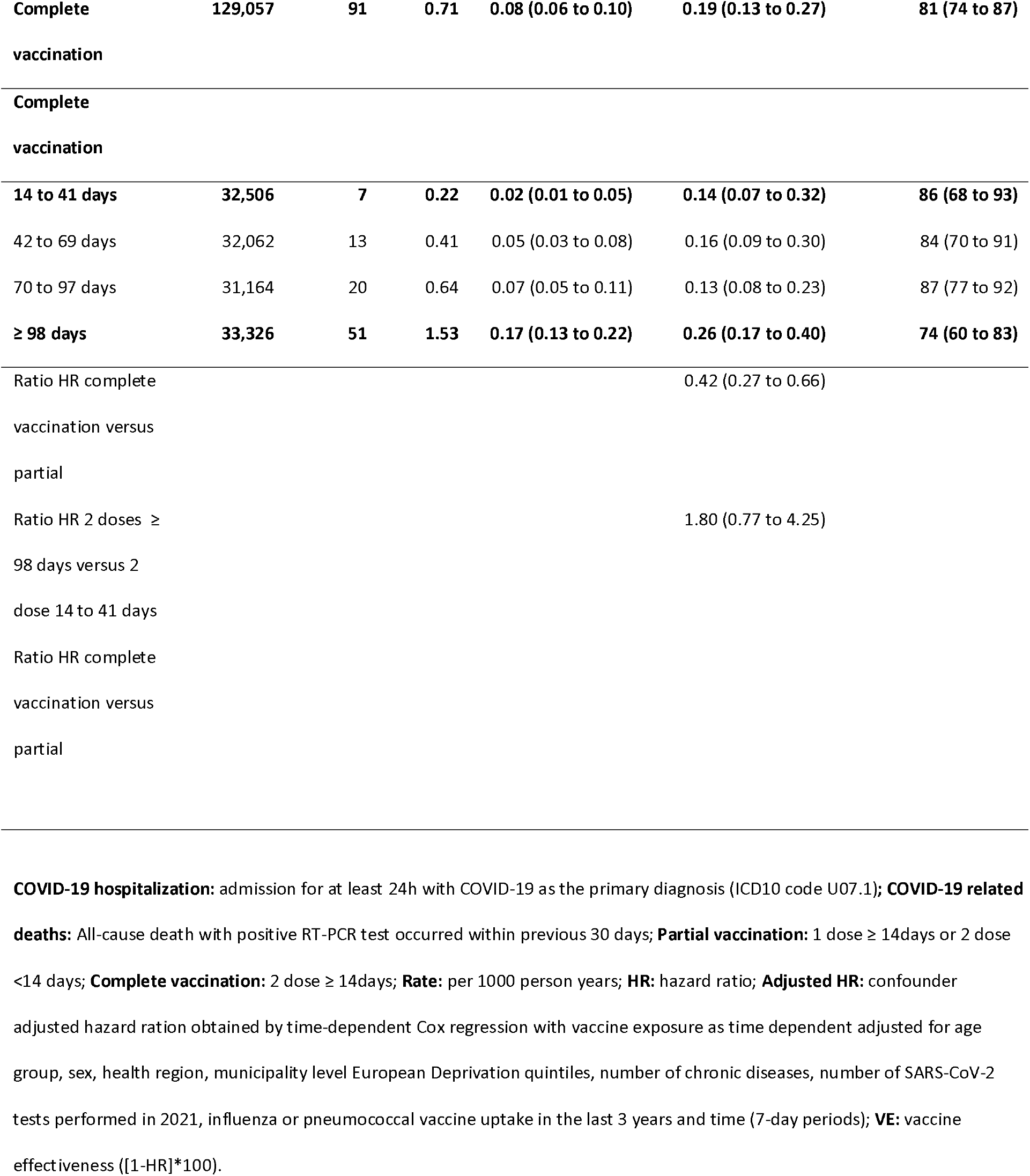
COVID-19 hospitalizations and COVID-19 related deaths, crude incidence rate (1000 person years), confounder-adjusted hazard ratios and vaccine effectiveness (%) by **number of mRNA doses and time in days since vaccine uptake, for individuals aged 80 years old or more**.

For both age groups, we observed a statistically significant increase of protection between partial and complete vaccination. Additionally, for the ≥80 yo cohort, we did not observe any statistical significant difference between VE estimates in persons with 98 or more days after the second dose (VE: 88%) compared with those with 14 to 41 days after the second dose (VE: 81%) (Table 4).

### Vaccine effectiveness against COVID-19 related deaths

Adjusted VE against COVID-19 related deaths, for the cohort of 65-79 yo, increased from 77% (95% CI 56 to 88) to 96% (95% CI 92 to 98) from partial to complete mRNA vaccination.

For the ≥80 yo VE against COVID-19 related deaths was 56% (95% CI 35 to 70) and 81% (95% CI 74 to 87), for partial and complete vaccinations, respectively. The risk of COVID-19 related deaths decreased significantly from partial to complete vaccination schemes in both age groups.

VE against COVID-19 related deaths, among those with 98 days or more after the second dose (VE=73%), was slightly lower than for those with 14 to 41 days (VE=86%) after the second dose, but the difference was not statistically significant (Table 4).

## Discussion

We present VE estimates of Pfizer-BioNTech or Moderna vaccines against severe COVID-19 outcomes for the Portuguese population aged 65 and older from February to August 2021. Our results indicate moderate to high protection against hospitalization with a primary diagnosis of COVID-19 (VE: 55-78%) and COVID-19 related deaths (VE: 56-77%) for partial mRNA vaccination and high levels of protection for the complete vaccination scheme (82-94% for hospitalization and 81-96% for mortality, respectively), supporting the advantage of complete vaccination.

For the complete vaccination scheme, our results for 65-79 yo are comparable to other studies conducted in Israel and the USA in population aged 65 and older, that reported Pfizer-BioNTech VE against hospitalizations of 97.9% and 94%, respectively [1,5].

The slight VE differences between our study and others may be explained by random variation, different study designs, observational periods and different epidemiological and virological contexts. Our study comprises a period of high COVID-19 incidence observed at the beginning of the vaccination campaign, corresponding to the third COVID-19 peak in January-February 2021 and a predominant Alpha VOC circulation, but also includes the period of its replacement by the SARS-CoV-2 Delta variant which in Portugal started in May 2021 [12].

mRNA VE estimates varied by age group for both severe outcomes targeted by this study, being about 23 percentage points (pp) lower for hospitalizations and 21 pp lower for COVID-19 related deaths for partial vaccination in the older cohort. For the complete vaccination, differences detected between age cohorts were smaller in magnitude, with 12 pp for hospitalizations and 15 pp for mortality.

Lower VE estimates observed in the older age cohort may be related to age-associated immunosenescence or waining of vaccine-induced protection, since ≥80 yo cohort in Portugal was targeted by the vaccination campaign earlier and had more prolonged exposure to the second dose. We measured VE by time since the second dose for the cohort ≥80 yo. Results suggest sustained VE levels up to 98 days (3 months) after the second dose for hospitalizations and a non-significant decrease for COVID-19 related deaths.

These results are consistent with recently published results on VE against hospitalization in the US general population [8]. Nevertheless, we cannot rule out bias in the VE estimates for the 98 days plus vaccine exposure class due to delayed data updates in hospitalizations and deaths.

The study has several limitations, some of which are related with the electronic registries used and their data quality. The main dataset used to link data was the NHSU, that contains the unique mandatory health number attributed to each individual in Portugal. However, as referred, the NHSU database could have update issues, and occasional register of NHS users, that could artificially increase the number of individuals in a given cohort. Several exclusion criteria were applied to overcome this limitation and the final cohort was comparable to the National Statistics Office estimates for individuals aged 65 and more years (Table S3) [13].

The delay of information on hospital discharge might contribute to underrepresenting this specific outcome and underestimating incidence rates for the more recent observation period. Although we did not find any reason for different delay in discharge information registry between vaccinated and unvaccinated, there is a possibility of a differential bias if vaccinated had a lower hospitalization time when compared to non-vaccinated. Our data do not allow us to estimate the length of hospitalization for each group and mitigate this potential bias.

Although we adjusted VE for calendar time, chronic conditions, and other relevant confounders, some residual confounding bias could be present mainly regarding the absence of information on personal protective measures like mask use, hand washing, and social distancing.

Finally, we were not able to estimate VE for other vaccine brands in 65-79 yo cohort due to their recent introduction and short follow up period, mainly after complete vaccination scheme.

In conclusion, our study supports high mRNA vaccine effectiveness for the prevention of COVID-19 hospitalizations and deaths in population with 65 or more years of age with a complete vaccination. Up 98 days (3 months) of second dose uptake, we did not found evidence of VE reduction. Use of cohort study designs based on nationwide health records linkage is a feasible and timely approach to monitor VE.

## Data Availability

The data used in this study are sensitive and will not be made publicly
available.

