## Supplementary material for "mRNA vaccines effectiveness against COVID-19 hospitalizations and deaths in older adults: a cohort study based on data-linkage of national health registries in Portugal"

Tables

Table S1 – Study period times (days) and date of last event for COVID-19 hospitalization and deaths - cohort 65-79 and 80 yo or older.

| | Start | End | Duration (days) |
|---|---|---|---|
| 65-79 Cohort | | | |
| Hospitalization | 30.03.2021 | 11.08.2021 | 134 |
| Deaths | 30.03.2021 | 09.08.2021 | 132 |
| 80 + Cohort | | | |
| Hospitalization | 02.02.2021 | 11.08.2021 | 190 |
| Deaths | 02.02.2021 | 07.08.2021 | 186 |

Table S2– Distribution of time (days) since second dose for cohort 65-79 and ≥80 yo.

| | Median | Interquartile Range (IQR) | Minimum-Maximum |
|---|---|---|---|
| 65-79 yo Cohort | 78 | 71 to 94 | 1 - 113 |
| ≥80 yo Cohort | 125 | 112 to 145 | 1 - 168 |

Table S3 – Comparison between the age group, sex and region distribution of the individuals included in the 65-79 age cohort and the Portuguese population estimates (Statistics Portugal, 2020 population estimates).

| | Cohort (%) | Population (%) |
|---|---|---|
| 65-79 yo cohort | | |
| Age group | | |
| 65-69 | 341,953 (38.9) | 602,210 (38.4) |
| 70-74 | 298,253 (34.0) | 536,388 (34.2) |
| 75-79 | 238,283 (27.1) | 428,706 (27.4) |
| Sex, male | 385,128 (43.8) | 694,974 (44.3) |
| ≥80 yo cohort | | |
| Age group | | |
| 80-84 | 232,429 (50.4) | 337797 (51.0) |
| 85+ | 228,391 (49.6) | 324027 (49.0) |
| Sex, male | 186,014 (40.4) | 239,436 (36.2) |

Figures

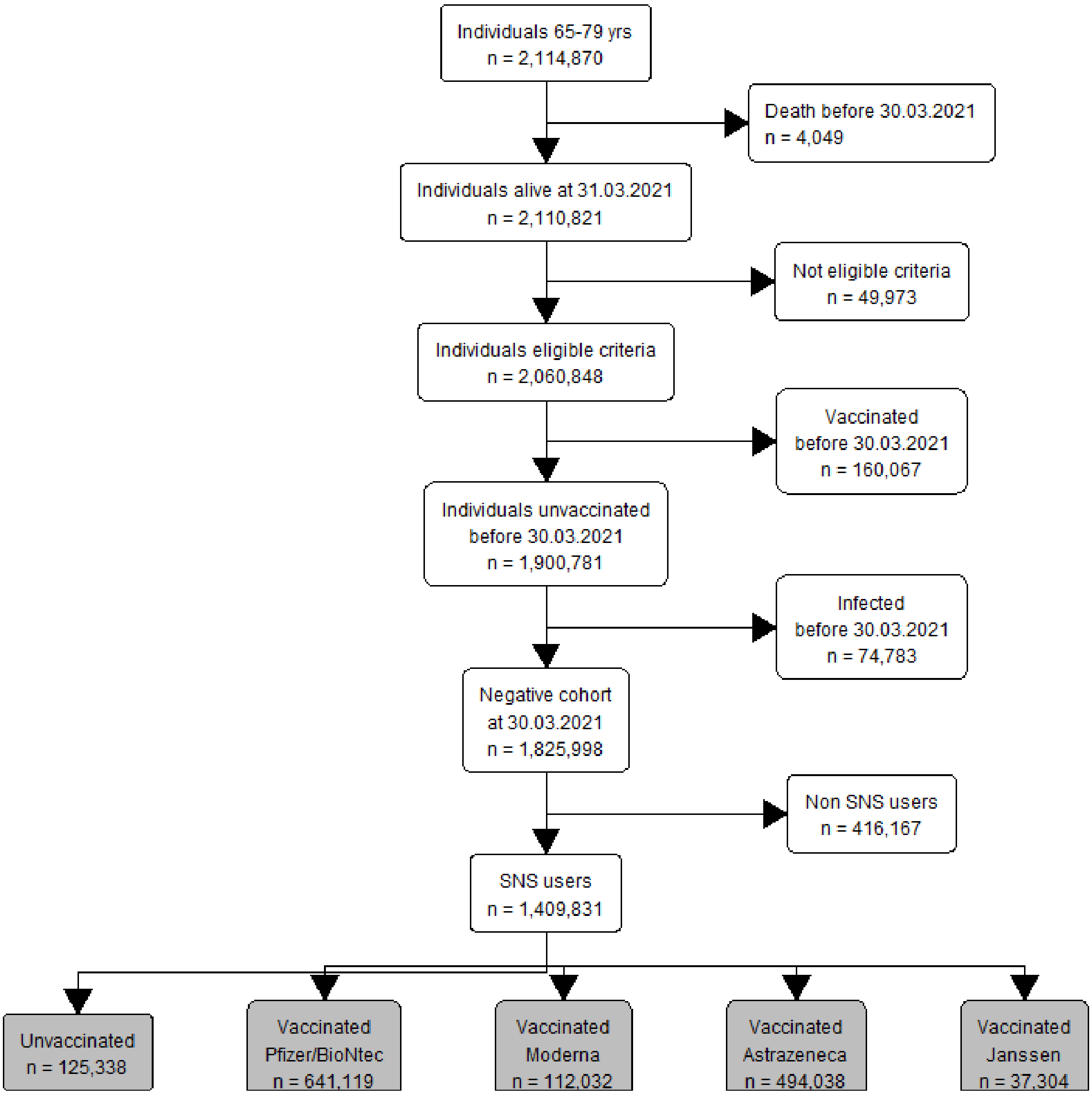


Figure S1 – Participants selection flowcharts 65-79 years old cohort

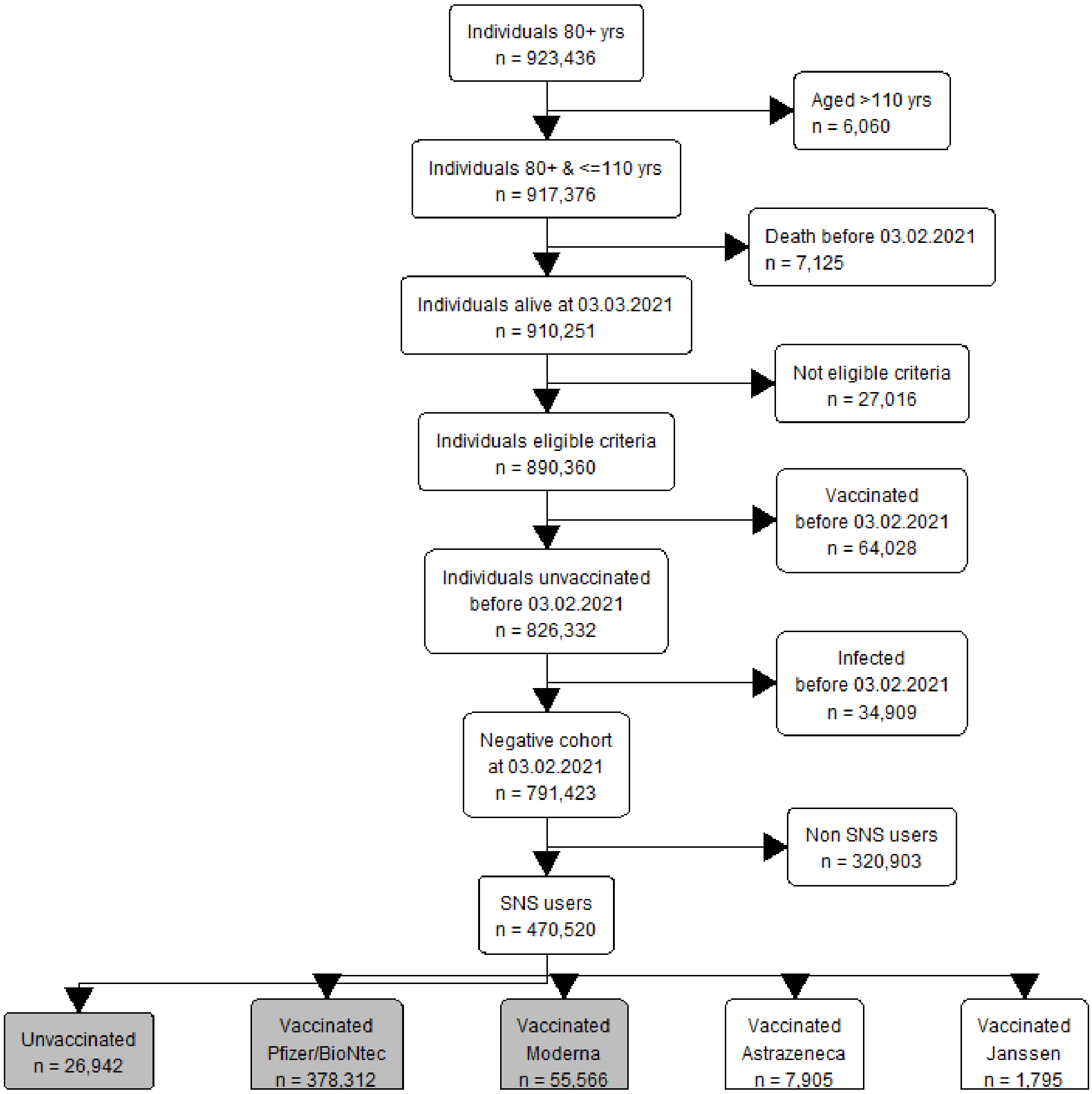


Figure S2 - Participants selection flowcharts ≥ 80 years old cohort

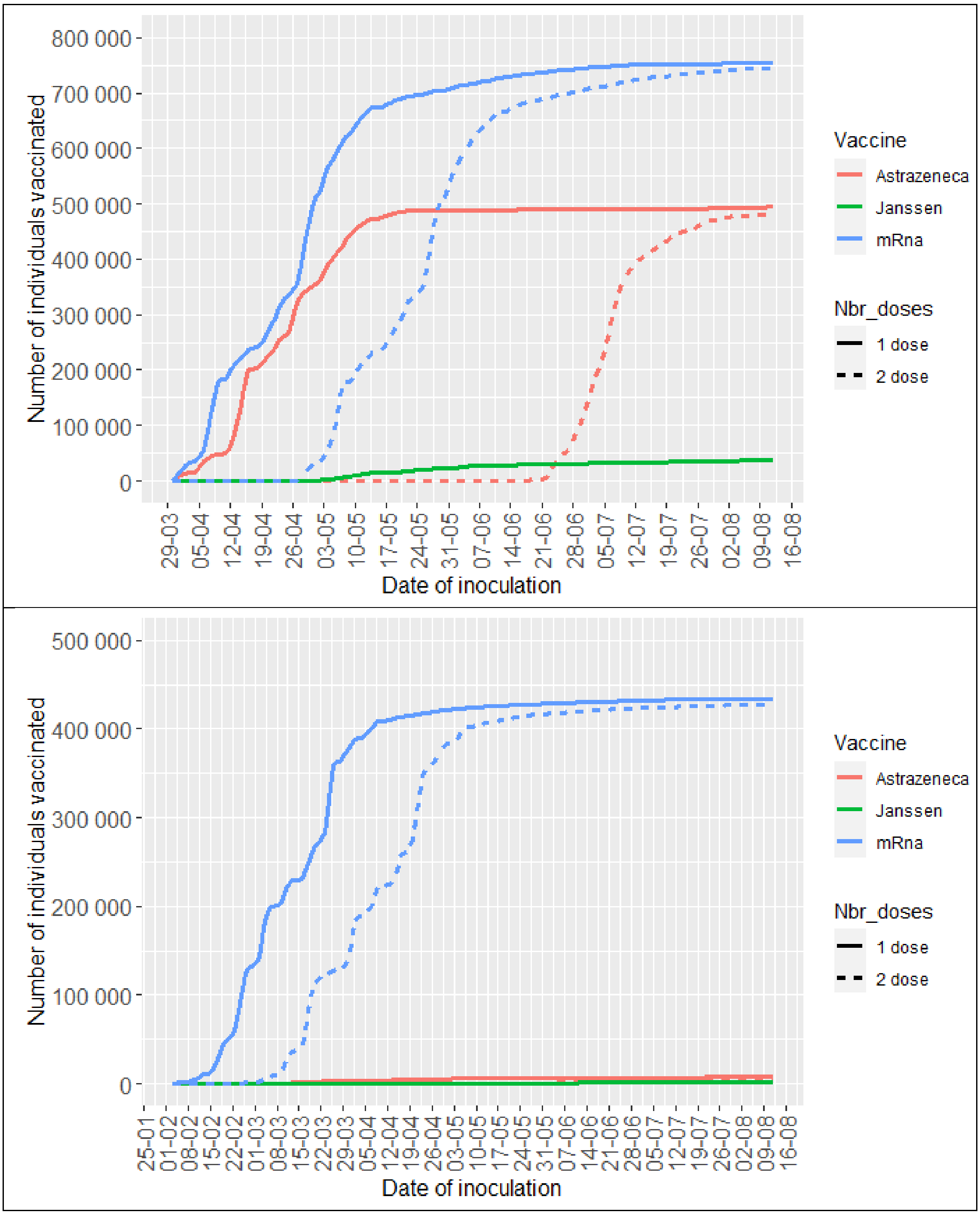


Figure S3– Daily accumulated number of individuals vaccinated with mRNA vaccines, according to the number of doses for the 65-79 age cohort (upper painel) and ≥80 years old cohort (lower painel).

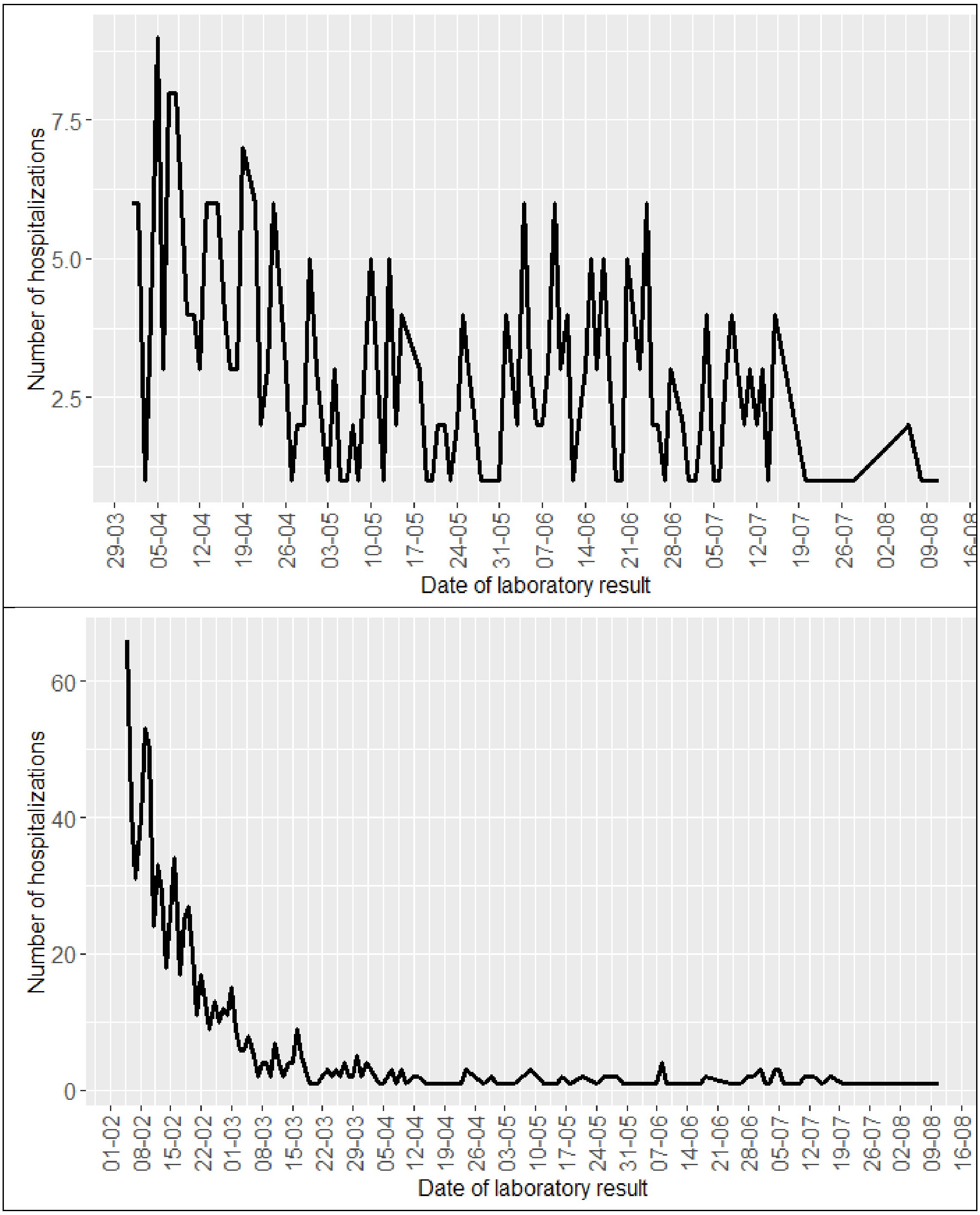


Figure S4 – Daily number of COVID-19 hospitalizations according to date of SARS-CoV-2 infection diagnosis for 65 to 79 cohort (upper painel) and ≥80 years old cohort (lower painel)

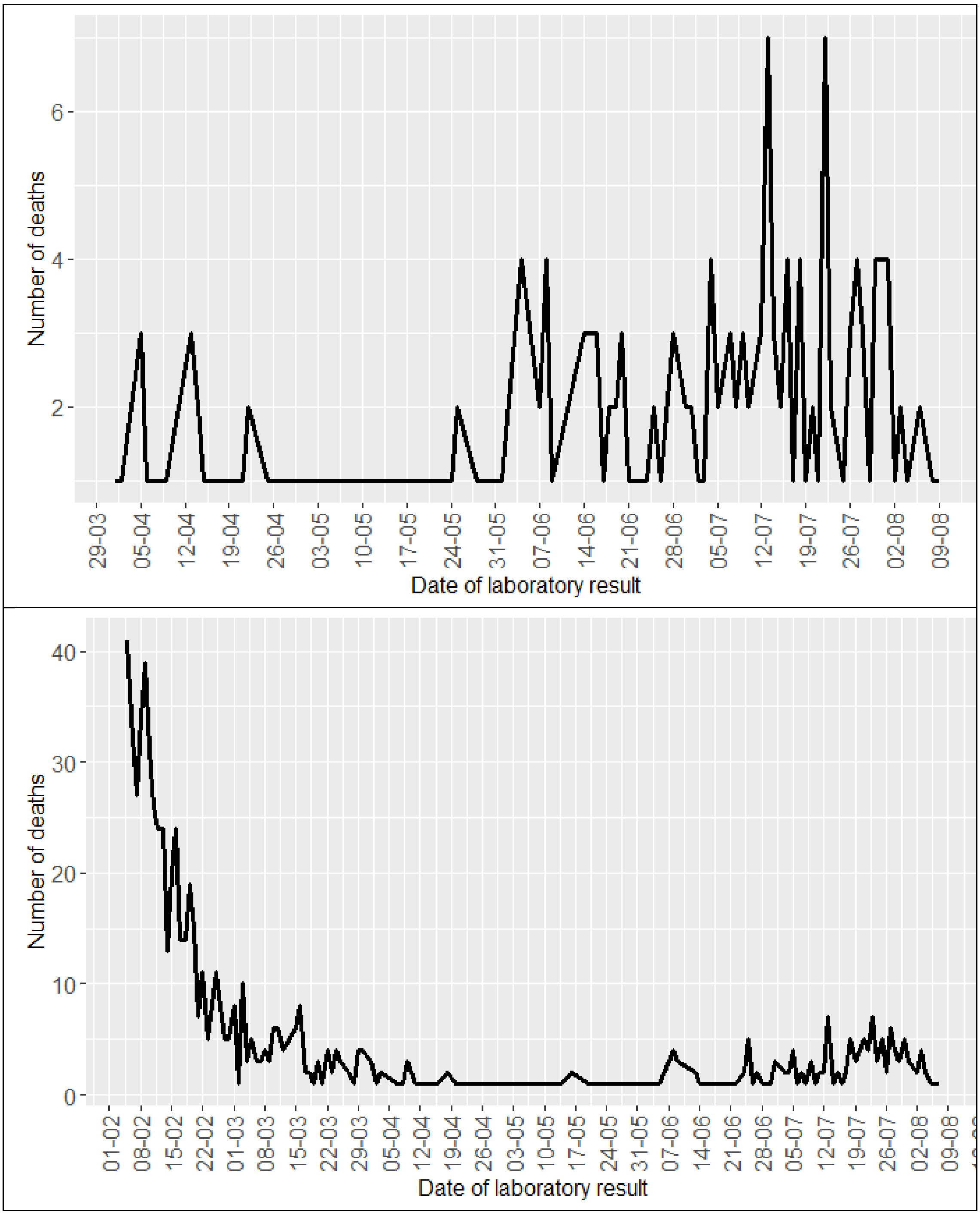


Figure S5 – Daily number of COVID-19 related deaths according to date SARS-CoV-2 infection diagnosis for 65 to 79 cohort (upper painel) and ≥80 years old (lower painel)
